# Mapping structural variants to rare disease genes using long-read whole genome sequencing and trait-relevant polygenic scores

**DOI:** 10.1101/2024.03.15.24304216

**Authors:** Cas LeMaster, Carl Schwendinger-Schreck, Bing Ge, Warren A. Cheung, Rebecca McLennan, Jeffrey J. Johnston, Tomi Pastinen, Craig Smail

## Abstract

Recent studies have revealed the pervasive landscape of rare structural variants (rSVs) present in human genomes. rSVs can have extreme effects on the expression of proximal genes and, in a rare disease context, have been implicated in patient cases where no diagnostic single nucleotide variant (SNV) was found. Approaches for integrating rSVs to date have focused on targeted approaches in known Mendelian rare disease genes. This approach is intractable for rare diseases with many causal loci or patients with complex, multi-phenotype syndromes. We hypothesized that integrating trait-relevant polygenic scores (PGS) would provide a substantial reduction in the number of candidate disease genes in which to assess rSV effects. We further implemented a method for ranking PGS genes to define a set of core/key genes where a rSV has the potential to exert relatively larger effects on disease risk. Among a subset of patients enrolled in the Genomic Answers for Kids (GA4K) rare disease program (N=497), we used PacBio HiFi long-read whole genome sequencing (lrWGS) to identify rSVs intersecting genes in trait-relevant PGSs. Illustrating our approach in Autism (N=54 cases), we identified 22,019 deletions, 2,041 duplications, 87,826 insertions, and 214 inversions overlapping putative core/key PGS genes. Additionally, by integrating genomic constraint annotations from gnomAD, we observed that rare duplications overlapping putative core/key PGS genes were frequently in higher constraint regions compared to controls (P = 1×10^−03^). This difference was not observed in the lowest-ranked gene set (P = 0.15). Overall, our study provides a framework for the annotation of long-read rSVs from lrWGS data and prioritization of disease-linked genomic regions for downstream functional validation of rSV impacts. To enable reuse by other researchers, we have made SV allele frequencies and gene associations freely available.

## INTRODUCTION

Structural variants (SVs) are a significant source of genetic diversity and have been increasingly recognized as contributors to rare and complex disease (Groza et al. 2024; Kainer et al. 2023; Merker et al. 2018). Typical SVs are genetic alterations longer than 50 bp in length and classified as deletions, duplications, insertions, and inversions. Considering such lengths, the analysis of SVs has been constrained by the limitations of short-read sequencing, which often fails to accurately resolve complex and repetitive genomic rearrangements (Merker et al. 2018; Marx 2023). In recent years, new long-read high fidelity (HiFi) sequencing technologies have enabled the detection and characterization of SVs with greater resolution and accuracy (Hon et al. 2020; Kucuk et al. 2023).

Despite improved SV detection, in a rare disease context it remains highly challenging to identify candidate pathogenic SVs. One solution to this challenge is to first understand the landscape of disease-associated genes for a given rare disorder. One such approach is in integrating trait-relevant polygenic scores (PGS), which quantify inherited genetic predisposition for a given phenotype by estimating disease effects across multiple genetic loci (Simona et al. 2023). In a rare disease context, increased polygenic liability has been observed for many disease cohorts, including in schizophrenia (Davies et al. 2020), severe neurodevelopmental disorders (Niemi et al. 2018), and autism spectrum disorder (Gaugler et al. 2014). Through integrating PGS, we can gain enhanced resolution into genes with impacts on rare diseases.

In this study, we leveraged PacBio long-read HiFi genomes available in the Genomic Answers for Kids (GA4K) study at Children’s Mercy Research Institute to investigate the role of rare structural variants (rSVs) in autism spectrum disorder. The GA4K study is a large-scale, phenotypically diverse pediatric rare disease cohort of over 13,000 individuals (Cohen et al. 2022). At the time of our study, there were 497 rare disease probands with HiFi long read whole genome sequences (lrWGS) completed. Further integrating open-source PGS available from PGS Catalog (Lambert et al. 2021), we computed individual PGS liability for autism spectrum disorder and additional traits and phenotypes. Our study focused on rare deletions, duplications, insertions, and inversions overlapping trait-relevant PGS genes. We further conducted a genomic constraint analysis of rSVs, providing insights on their evolutionary tolerance and potential pathogenicity.

Overall, we found an enrichment of deletions, duplications and insertions at trait-relevant genes and a significantly higher constraint of duplications in individuals with autism. Furthermore, we trace SV-overlapped genes back to autism and implicate anterograde trans-synaptic signaling using ontology enrichment. Our study utilizes the intersection of rSVs, trait-relevant PGS, and genomic constraint to explore the structural variant landscape in a large-scale rare disease cohort, with a focus on genes associated with autism spectrum disorder.

## RESULTS

### Structural variant type and frequency show prevalence of deletions and insertions

We analyzed 497 probands with HiFi long read whole genome sequences (lrWGS) in the Children’s Mercy GA4K database (N = 254 male; N = 243 female). We assessed ancestry using Somalier, which predicted 437 European, 58 Admixed American, and 2 Asian individuals.

Structural variants (SVs) were called using PacBio SV (PBSV) after alignment with human genome GrCh38 (Pedersen et al. 2020). All individuals were assessed for total number of SVs (**Supplemental Table 1, Supplemental Figure 1**). We developed a python application called *GA4K-SV-FINDER* where researchers can query these SVs, including allelic frequencies and overlapping genes, by inputting user-specified genomic regions or gene names (**Data Availability**).

To identify potential large-effect rare structural variants (rSVs) impacting rare disease phenotypes we removed common SVs from the dataset (>0.5% MAF; **Methods**) (**Supplemental Figure 1**). This reduced a total of 10,631,806 SVs to 1,017,180 rSVs (**Figure 1A**). The mean number of SVs per individual was 21,521, and 2,047 for rSVs (**Figure 1A**).

**Figure 1:**
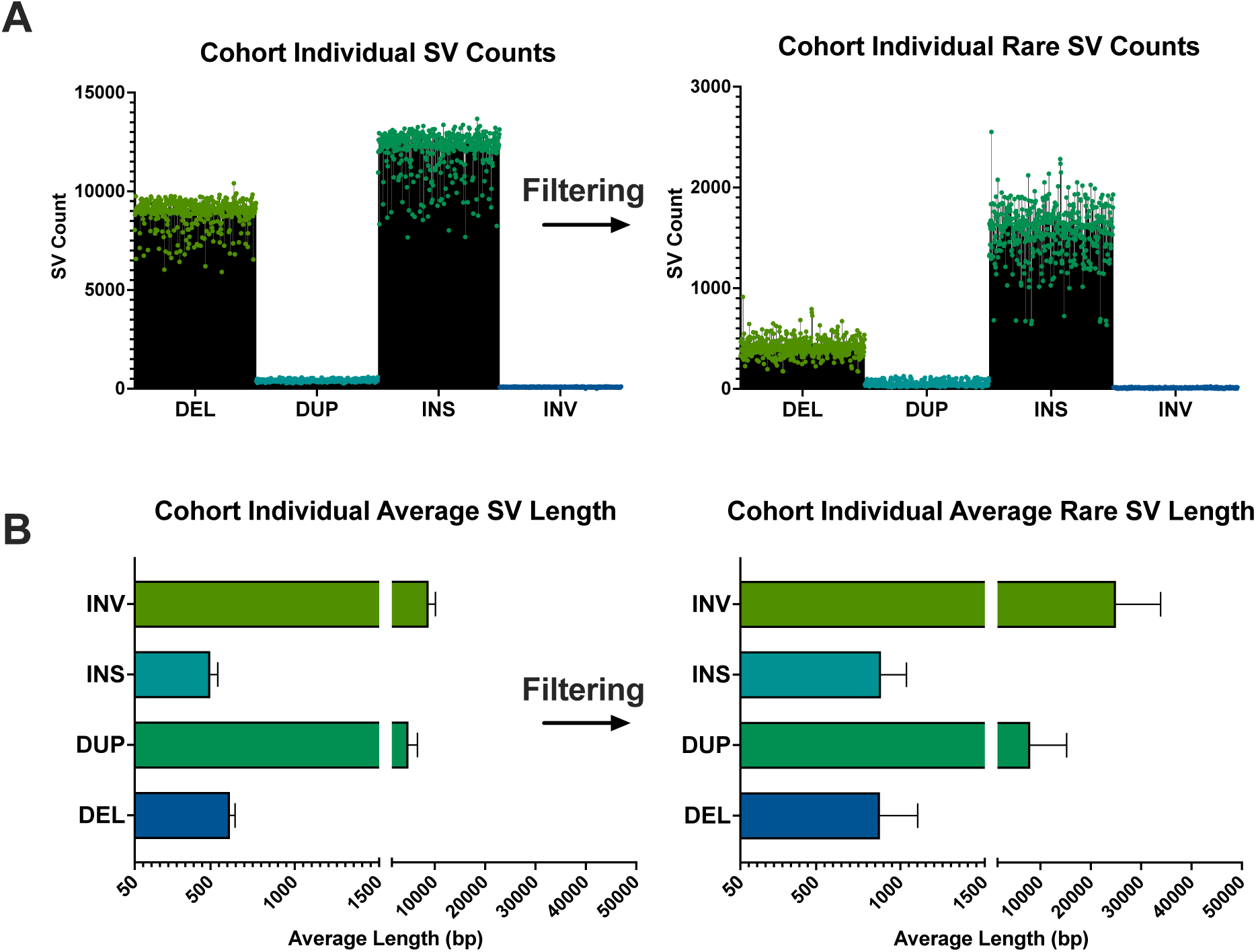
SV type frequency and average length in study population. (**A**) Two dot-bar graphs representing SV type (Deletions (DEL), duplications (DUP), insertions (INS), inversions (INV)) counts before and after filtering for rare variants. (**B**) Two horizontal bar graphs with outlier whiskers displaying the average lengths for SVs before and after filtering.

Representing the lowest frequency (0.5%) of rSV type, inversions (INVs) had the largest mean length (25,022 bp) compared to deletions (DEL, 20.3%, 879 bp), duplications (DUP, 2.4%, 8,000 bp), and insertions (INS, 76.8%, 885 bp) (**Figure 1**). When comparing SV type across sexes, frequencies were similar and within 5% variability between males and females for length and count. The frequency and length of rSVs in our study closely correspond with population frequencies seen in other studies (Kosugi et al. 2019; Guo et al. 2021).

### Cohort phenotypes aligned with polygenic scores in five GA4K phenotypes

We calculated PGS for all probands across various rare phenotypes (short stature/height; global developmental delay/fluid intelligence; autism/autism; hypotonia/grip strength; and seizure/epilepsy). Probands were then grouped by their phenotype to construct case/control cohorts. Probands not possessing a given phenotype were considered controls in each mapping. We chose to analyze the top-five most frequent phenotypes in GA4K that had accompanying lrWGS. At the time of assessment, the largest phenotype was Global Developmental Delay at 138 cases, followed by Seizures (86), Hypotonia (62), Autism (54), and Short Stature (45) (**Figure 2A**). We observed significant differences in Autism and Short Stature PGS, with scores higher for Autism and lower for height, respectively (**Figure 2A**).

**Figure 2:**
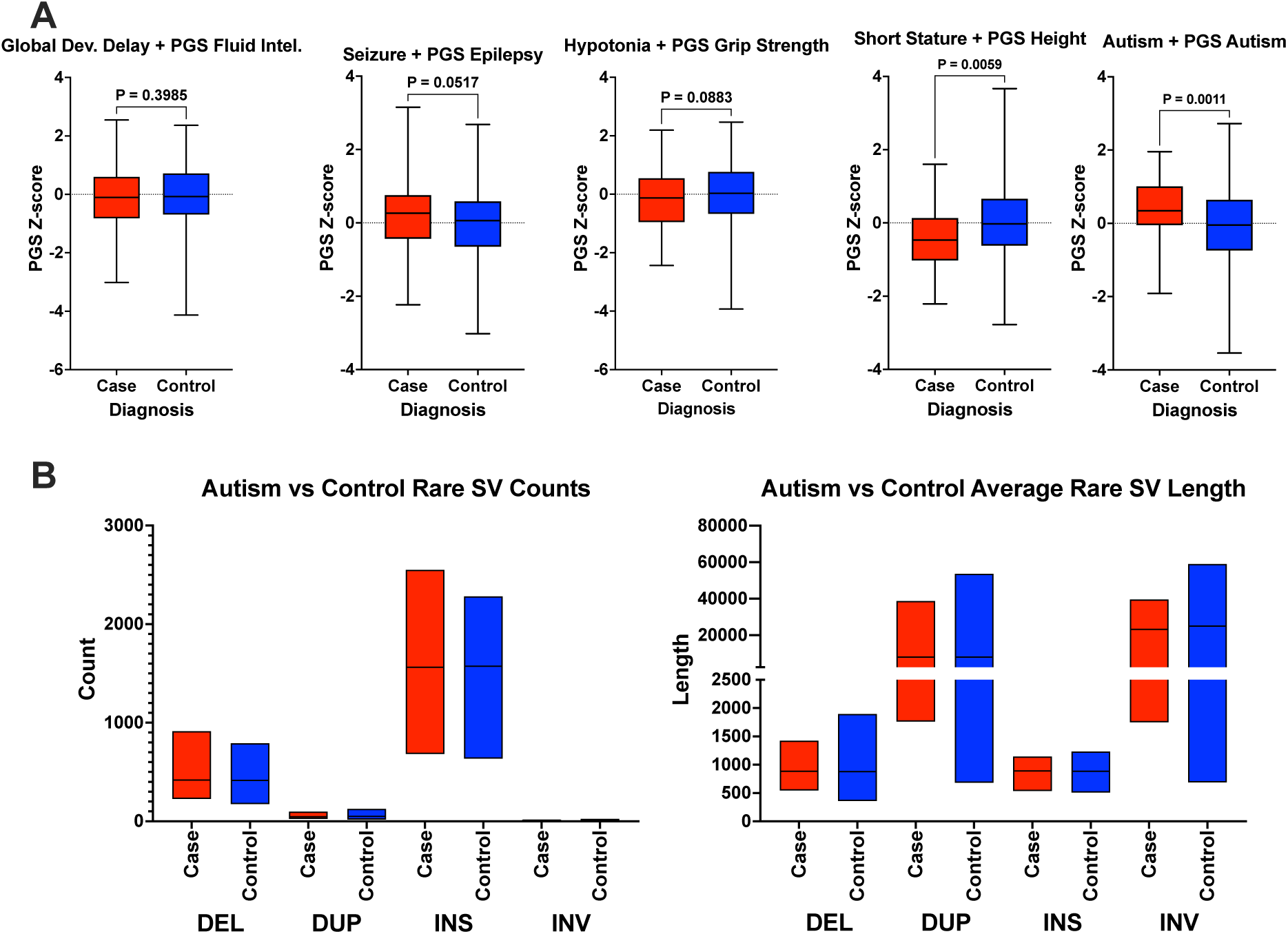
Box plots show significant mappings for Autism and Short Stature. (**A**) Box and whisker plots showing differences in selected PGS mapped to five rare disease phenotypes in GA4K for cases and controls. Red indicates case individuals. Blue indicates control individuals. PGS Z-score is indicated on the Y axis (N cohort = 497, N GDD cases = 138, N Seizure = 86, N Hypotonia = 62, N Autism = 54, N Short Stature = 45). P-values were computed using a Mann-Whitney test. (**B**) Box plots showing the count of rSV type (DEL, DUP, INS, INV) (left) and rSV type length (right) across individuals for cases (N = 54) and controls (N = 443) in the Autism cohort.

Focusing on the larger case number of the two significant phenotypes, we then compared rSVs in the Autism group (N cases = 54) group with controls (N controls = 443). We observed that probands with Autism had slightly higher median counts of DUPs and INVs, and longer mean lengths, except for INVs, when compared with controls (**Figure 2B**).

### High weight PGS genes reveal rSV enrichment in Autism

Due to the complex genetic nature of Autistic disorders, we decided to further focus on individuals in our cohort with Autism. We evaluated genes harboring variants included in a PGS for Autism (PGS Catalog ID: PGS000327). Each variant is weighted according to its relative association with a phenotype (**Methods**). We intersected the PGS variant coordinates with the coordinates of genes. This revealed 5,830 genes intersected by at least one overlapping PGS variant. We then intersected cohort rSVs with these PGS-linked genes and found at least one overlap in 4,271 genes (**Supplemental Table 2**). Among this gene set, Ras Suppressor Protein 1 (RSU1) had the highest PGS weighting (gene weight = 0.139). The lowest weighted gene was Importin 7 (IPO7) (gene weight = 0.023) (**Supplemental Table 2; Figure 3**). Additionally, there are 262 genes associated with the Human Phenotype Ontology (HPO) ID for autism spectrum disorder (HP:0000717) (Kohler et al. 2021); of those genes, 84 were found to have overlap with rSVs (average gene weight = 0.05).

**Figure 3:**
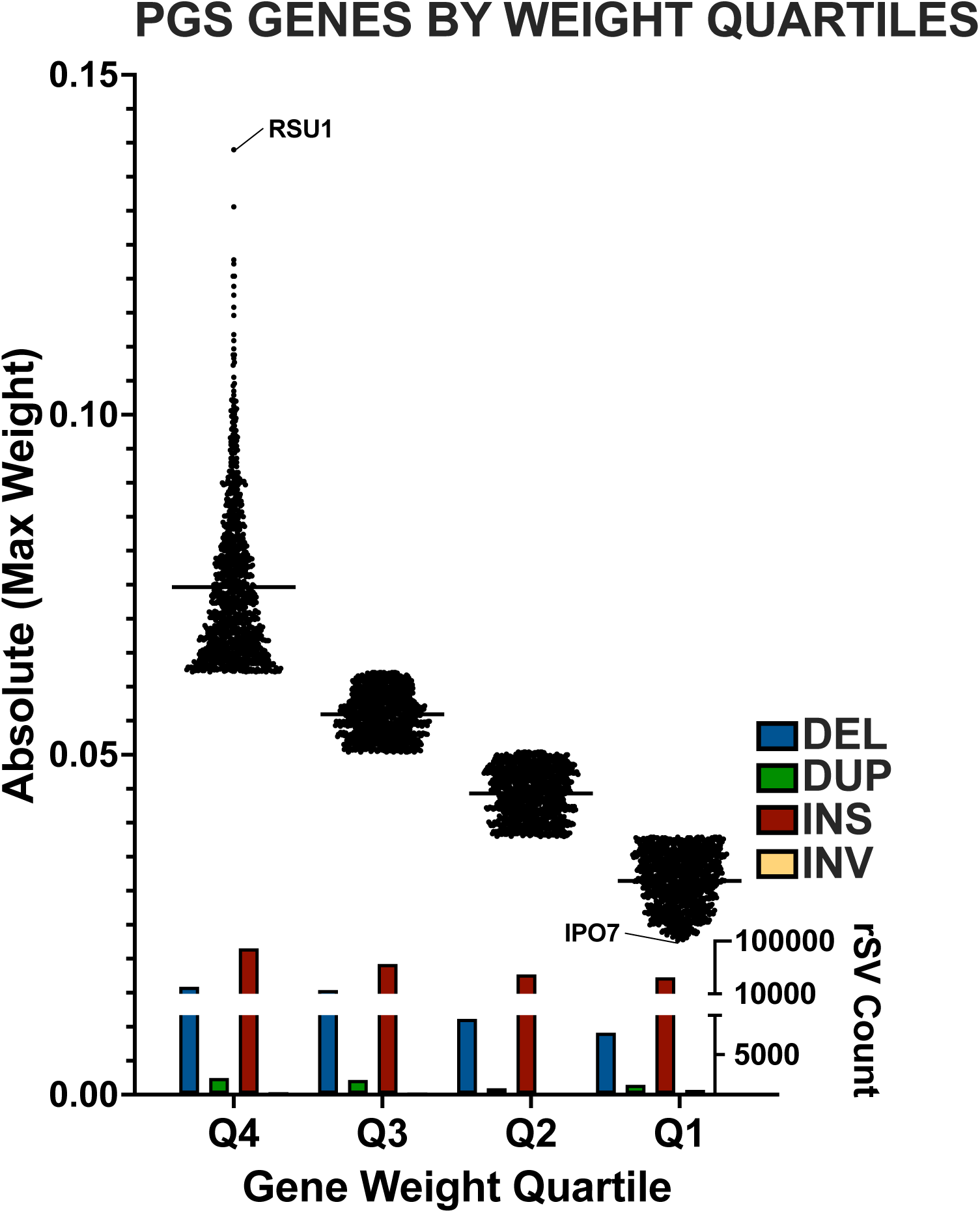
Top quartile PGS genes show a wide range of weights and a high count of insertions. Dot plot of genes within their quartiles based on the intersecting PGS gene weights. The highest weighted genes are in Q4 on the left. The lowest weighted genes are in Q1, to the right. The y-axis for gene weights displays the range of gene weights. The inset bar plot shows the rSV count for each quartile. A color legend is displayed above the y-axis.

We then organized these 4,271 genes into 4 quartiles according to their PGS gene weight (**Figure 3**). Each quartile contained 1063-1071 genes. All 4,271 genes were evaluated for frequency of rSVs both within the gene body and 200kb up- and down-stream of the gene body (**Supplemental Table 2**). Focusing on the top quartile, we observed 112,100 rSV overlaps. INSs were the most abundant, both in the gene body (78.3%) and in the flanking regions (upstream: 77%, downstream: 79.3%). This was followed by DELs (body: 19.6%, up: 20.5%, down: 18.1%), DUPs (body: 1.8%, up: 2.1%, down: 2.4%), and INVs (body: 0.2%, up: 0.3%, down: 0.2%). INS and DEL frequency in the gene body paralleled a descending gene weight across all quartiles, where lower gene weights correlated with a lower number of the SV (**Figure 3**). Duplications followed a similar trend, but the bottom quartile had more than the third quartile. Inversions were more abundant in the bottom quartile, however 431 of 514 INVs were found overlapping the gene Chymotrypsinogen (CTRB1/CTRB2) – a gene region harboring many INVs as previously described (Rosendahl et al. 2018). When evaluating the up- and down-stream 200kb flanking regions for each gene, higher up- and down-stream rSV frequencies associated with a lower gene weight quartile, where the lower gene weight had higher flanking rSV counts.

We then quantified genes in the top quartile of PGS effect with rSV frequency significantly different (>5% diff, Chi-Square test, p-value < 0.05) between Autism and control groups (**Table 1; Methods**). In doing this we move from entire cohort frequencies to an Autism-control enrichment. While we define significant frequency differences between groups where genes have >5% enrichment over the other group, our aim here is to resolve higher impact rSV-gene enrichments between Autism and controls (>10%) in the top quartile for in-depth downstream analysis (**Table 1**). Following this approach, we observed that DELs were enriched in GRNA Binding Motif Single Stranded Interacting Protein 3 (RBMS3), Contactin-associated Protein 2 (CNTNAP2), Human Immunodeficiency Virus Type I Enhancer Binding Protein 3 (HIVEP3), Ribosomal Protein S6 Kinase A5 (RPS6KA5), Myomesin 2 (MYOM2), and Astrotactin-2 (ASTN2) (**Table 1**). Only one gene, Repeat-Containing Protein 7 (WDR7), had >10% enrichment of DUPs (**Table 1**). The most frequent rSV type, INSs, was enriched in Catenin Alpha-3 (CTNNA3), Neuronal Guanine Nucleotide Exchange Factor (NGEF), Calmodulin-lysine N-methyltransferase (CAMKMT), Coiled-Coil Domain Containing 171 (CCDC171), IQ Motif Containing GTPase Activating Protein 2 (IQGAP2), Myosin ID (MYO1D), Death-associated Protein Kinase 1 (DAPK1), and Neuregulin 3 (NRG3) (**Table 1**). No INVs were found to be > 10% differentially enriched (**Table 1**). The genes RBMS3, CTNAP2, HIVEP3, ASTN2, CTNNA3, and DAPK1 have been previously reported in connection to autism spectrum disorders (Pereanu et al. 2018).

**Table 1:**
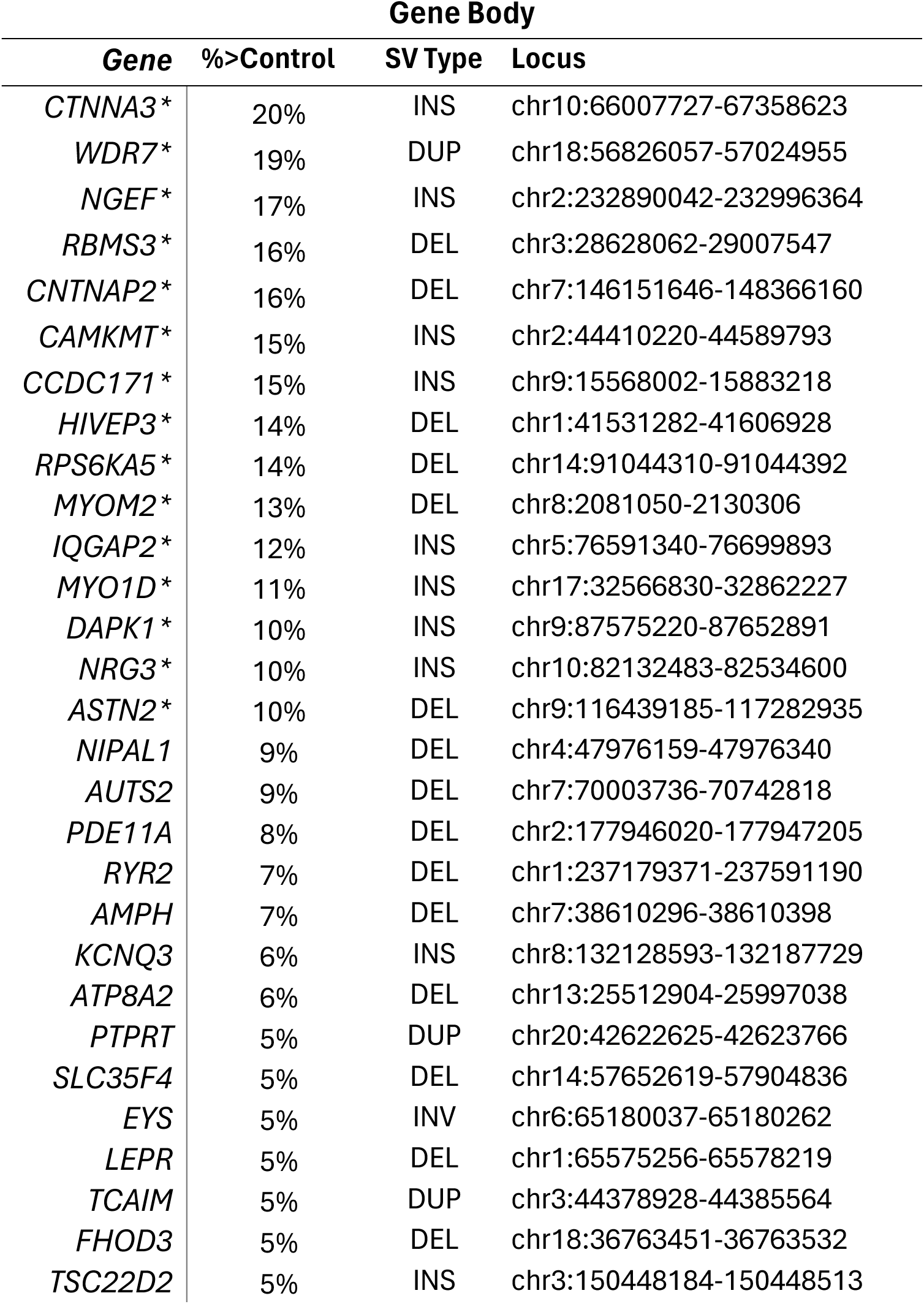
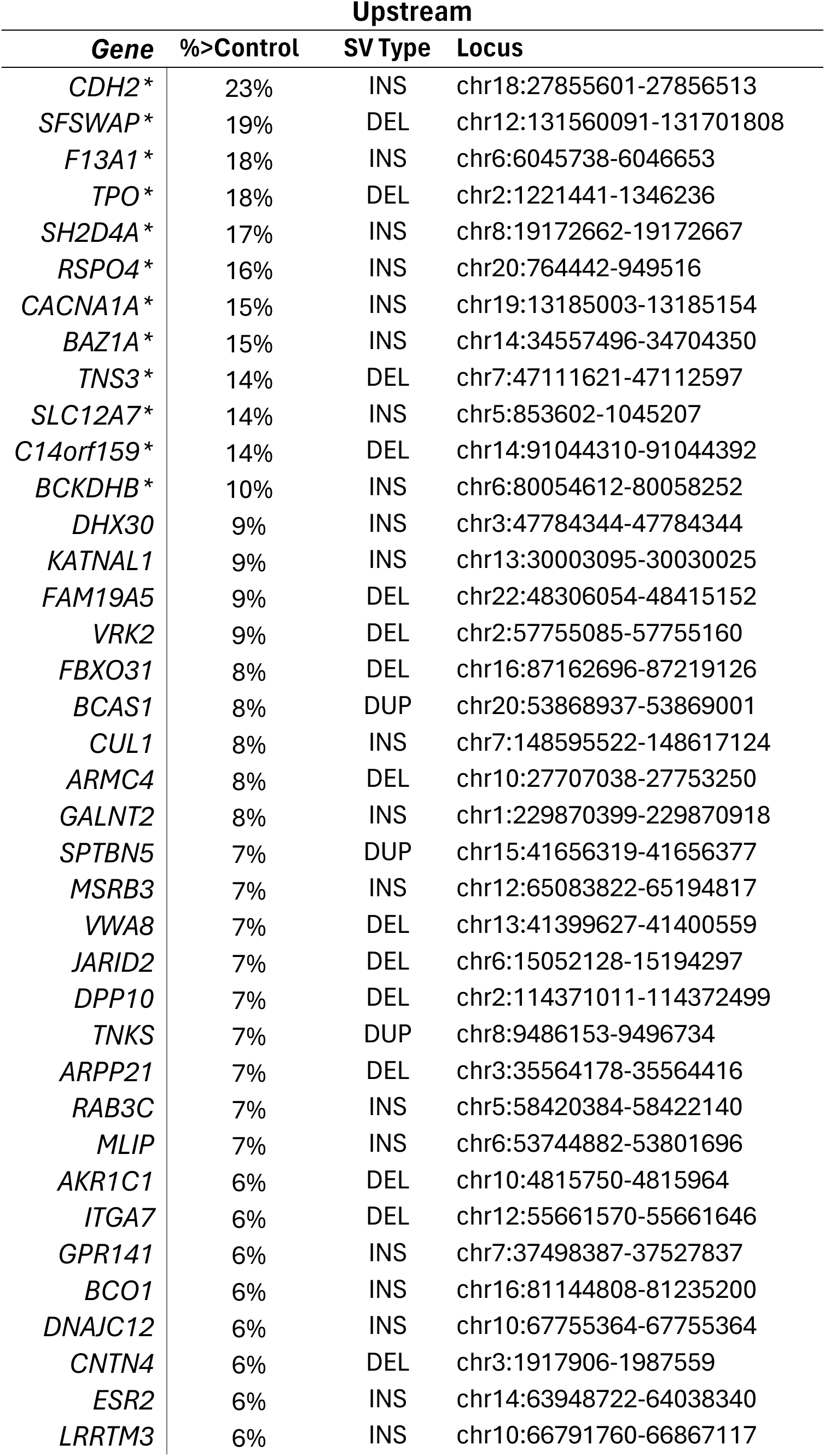
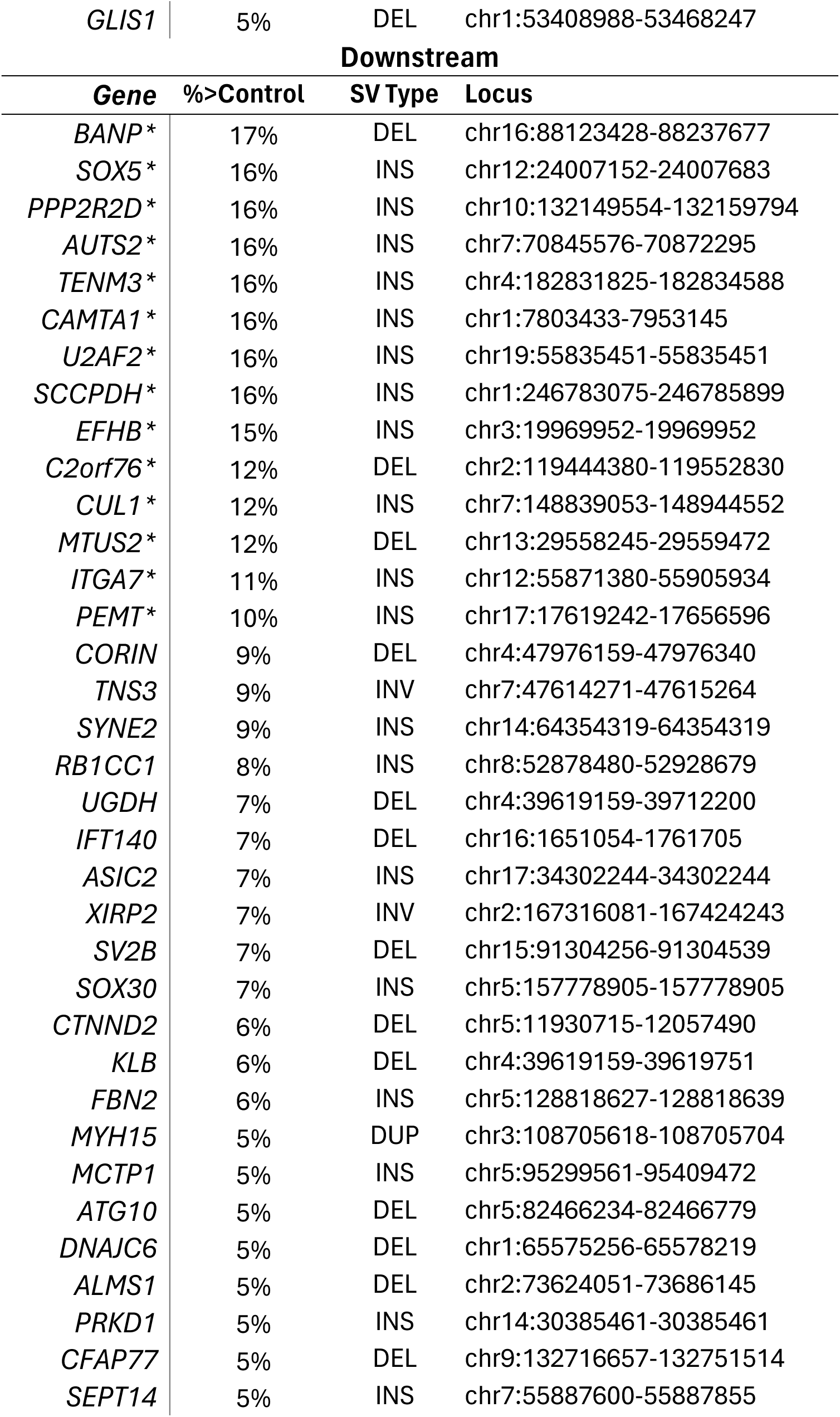
SV enriched genes strongly associated with Autism. Genes with significantly (Chi-Square, p-value < 0.05) higher frequency rSV enrichment (>5%) in Autism individual gene bodies (top), upstream (middle) and downstream (bottom) regions. The group frequency differential and SV type is displayed to the right of the gene. *Indicates high impact (>10%) rSV-gene differential enrichment in Autism over controls.

We also assessed enrichment in the 200kb flanking regions of these high weight PGS-linked genes, revealing significant enrichment of DELs down-stream of BTG3 Associated Nuclear Protein (BANP), MOB Kinase Activator 1A (C2orf6), and Microtubule Associated Scaffold Protein 2 (MTUS2). However, most down-stream enrichments were INSs proximal to SRY-box Transcription Factor 5 (SOX5), Protein Phosphatase 2 Regulatory Subunit B Delta (PPP2R2D), Autism Susceptibility Candidate 2 (AUTS2), Saccharopine Dehydrogenase (SCCPDH), Teneurin Transmembrane Protein 3 (TENM3), Calmodulin Binding Transcription Activator 1 (CAMTA1), U2 Small Nuclear RNA Auxiliary Factor 2 (U2AF2), EF-Hand Domain Family Member B (EFHB), Cullin 1 (CUL1), Integrin Subunit Alpha 7 (ITGA7), and Phosphatidylethanolamine N-Methyltransferase (PEMT) (**Table 1**). Furthermore, we observed an enrichment of DELs up-stream of Splicing Factor SWAP (SFSWAP), Thyroid Peroxidase (TPO), Tensin 3 (TNS3), and D-Glutamate Cyclase (C14orf159), as well as INSs enriched up-stream of genes Cadherin 2 (CDH2), Coagulation Factor XIII A Chain (F13A1), SH2 Domain Containing 4A (SH2D4A), R-Spondin 4 (RSPO4), Calcium Voltage-Gated Channel Subunit Alpha1 A (CACNA1A), Bromodomain Adjacent To Zinc Finger Domain 1A (BAZ1A), Solute Carrier Family 12 Member 7 (SLC12A7), and Branched Chain Keto Acid Dehydrogenase E1 Subunit Beta (BCKDHB). We did not observe an enrichment for DUPs or INVs within the PGS-linked gene set.

While we focused on differential enrichment between groups, it is also important to note that the two genes with the highest frequency of rSVs in Autism individuals were not found in SFARI or AutDB Autism databases (MYOM2, 100% [state what this percentage means, just to remind the reader]; NGEF, 98%) (**Table 1**; **Figure 4**). Moreover, the top 3 most frequent genes associated with Autism – CTNNA3 (43%), CNTNAP2 (35%), and RBMS3 (26%), while differentially enriched in Autism individuals, were less frequent than those genes with previous associations to the disease (**Table 1**; **Figure 4**) (Pereanu et al. 2018). The only gene with enrichments in the gene body as well as a flanking region (downstream) was AUTS2, where DELs were found in the gene body in 13% of individuals and 42% had INSs in the downstream region (**Figure 4**). Furthermore, genes TNS3 (Upstream: 22% DEL; Downstream: 13% INV), CUL1 (Upstream: 11% INS; Downstream: 26% INS), and ITGA7 (Upstream: 7% DEL; Downstream: 24% INS) had enrichments in both up- and down-stream regions (**Figure 4**).

**Figure 4:**
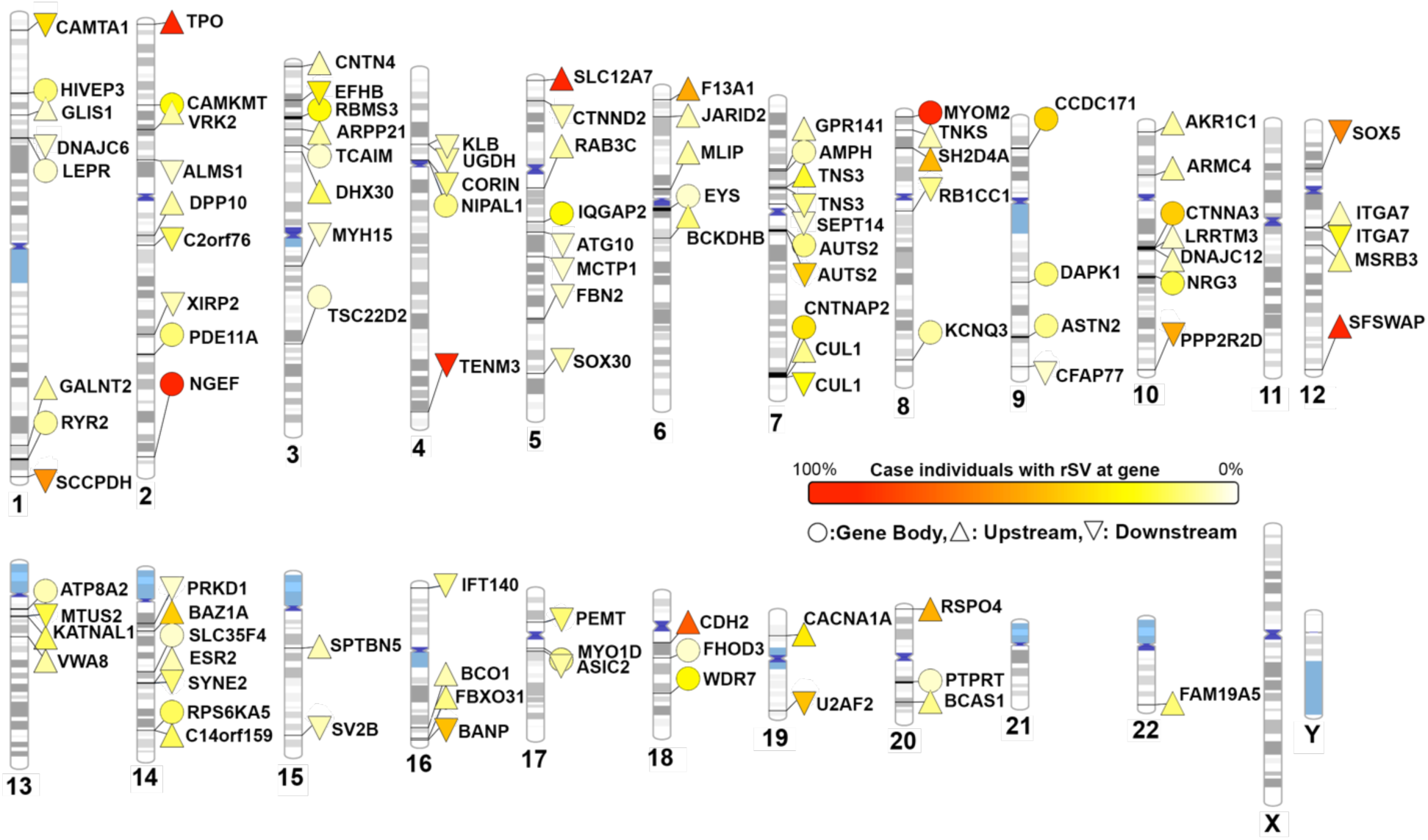
Genes with differential rSV enrichment tend have close chromosomal proximity to one another whereas rSV enriched genes with high frequency tend to have more distance between. PhenoGram showing the frequency of individuals with rSVs at the cut-off genes by chromosomal position (color scale from 0-100% frequency of case individuals). Cytogenic Giemsa bands show GC-(blue) and AT-rich (gray) regions. Symbols indicate whether the enrichment occurs in the gene body, the upstream, or the downstream regions of the listed gene. The color gradient of symbols indicates the degree of enrichment in the case cohort, with higher enrichment indicated in orange and red.

Insertions were the most common type of enrichment (49/103), followed by DELs (44/103), DUPs (7/103), and INVs (3/103). Two of the 19 chromosomes with rSV enrichments at high weight PGS-linked genes were exclusive to INSs. We also found that while DUPs were relatively infrequent, differentially enriching the gene body of 3 of 62 total genes, DUPs at WDR7 had the second highest differential percentage of an enriched gene body (19% > Controls, p value = 0.0001) (**Table 1**). Though WDR7 was not found in AutDB or in HPO gene associations, it is positioned down-stream of CDH2 on chromosome 18, the most frequently up-stream enriched gene (76%) associated with Autism (Abrahams et al. 2013). Additionally, PTPRT and BCAS1, two more of the seven genes enriched by DUPs, are both positioned on chromosome 20 and associated with Autism. Moreover, enriching DUPs were at genes with a mean gene weight in the top 3% of PGS-linked genes, whereas DELs, INSs, and INVs were in the top 14%. Overall, we identified genes with differential SV enrichments in Autism over control individuals, specific to the rSV type and locus, finding a high frequency enrichment of DELs and INSs overlapping PGS-linked genes and their flanking regions.

### SV enriched genes in the top quartile of PGS effect size significantly associate with autism and potentially associated pathways

After deriving a list of genes with high enrichment of rSVs in Autism (>5%; N genes = 103) (**Table 1**), we sought to determine if these genes specifically enrich the Autism phenotype or candidate biological pathways. We used the DisGeNET database with Enrichr and found a significant enrichment (adj. P = 5×10^−06^) with Autistic Disorder using our gene list (**Table 1**). Furthermore, using the GO Cellular Component database, we found that rSVs were enriched in cases for the genes Formin Homology 2 Domain Containing 3 (FHOD3), MYO1D, Katanin Catalytic Subunit A1 Like 1 (KATNAL1), AUTS2, DAPK1, MTUS2, Amphiphysin (AMPH), IQGAP2, CORIN, and Spectrin Repeat Containing Nuclear Envelope Protein 2 (SYNE2) all of which are involved in the cytoskeleton (GO:0015629; GO:0005856) (adj. p-value = 0.02) (**Table 1**). Our results here provide a framework for linking increased rSV burden in Autism to potential downstream impacts on gene pathways relevant to Autism pathogenesis.

### Duplications are enriched at regions of high constraint in top PGS genes

We next used 1kb gnomAD constraint windows to evaluate potential pathogenicity amongst rSVs (**Methods**). In short, higher constraint indicates an evolutionary intolerance to variation within that locus. Though constraint scores were not available across every 1kb region of the genome, we were able to intersect constraint windows for 7.9% of rSVs in the gene body and 6.7% in the flanking regions. We graphed the density of rSV constraint scores across different SV types (**Figure 5**). This analysis revealed that rare DELs and DUPs in the Autism group tended to overlap higher constraint windows compared with controls (DEL: p < 0.0001, Autism z mean = 0.14, control z mean = −0.08; DUP: p < 0.0001, Autism z mean = 0.48, control z mean = 0.03). Significant differences were not observed for INSs or INVs (INS: p = 0.66, Autism z mean = 0.2, control z mean = 0.18; INV: p = 0.14, Autism z mean = −2.77, control z mean = −2.6). Furthermore, Autism DUPs tended to overlap higher constraint windows within 200kb up-stream of PGS-linked genes (DUP: p < 0.0001, Autism z mean = 0.66, control z mean = 0.14). We observed a similar pattern for DUPs in Autism individuals for rSVs within the down-stream region (DUP: p < 0.0001, Autism z mean = 0.42, control z mean = 0.11). Thus, DUPs were the only SV type in Autism with significantly higher constraint than controls in all three regions.

**Figure 5:**
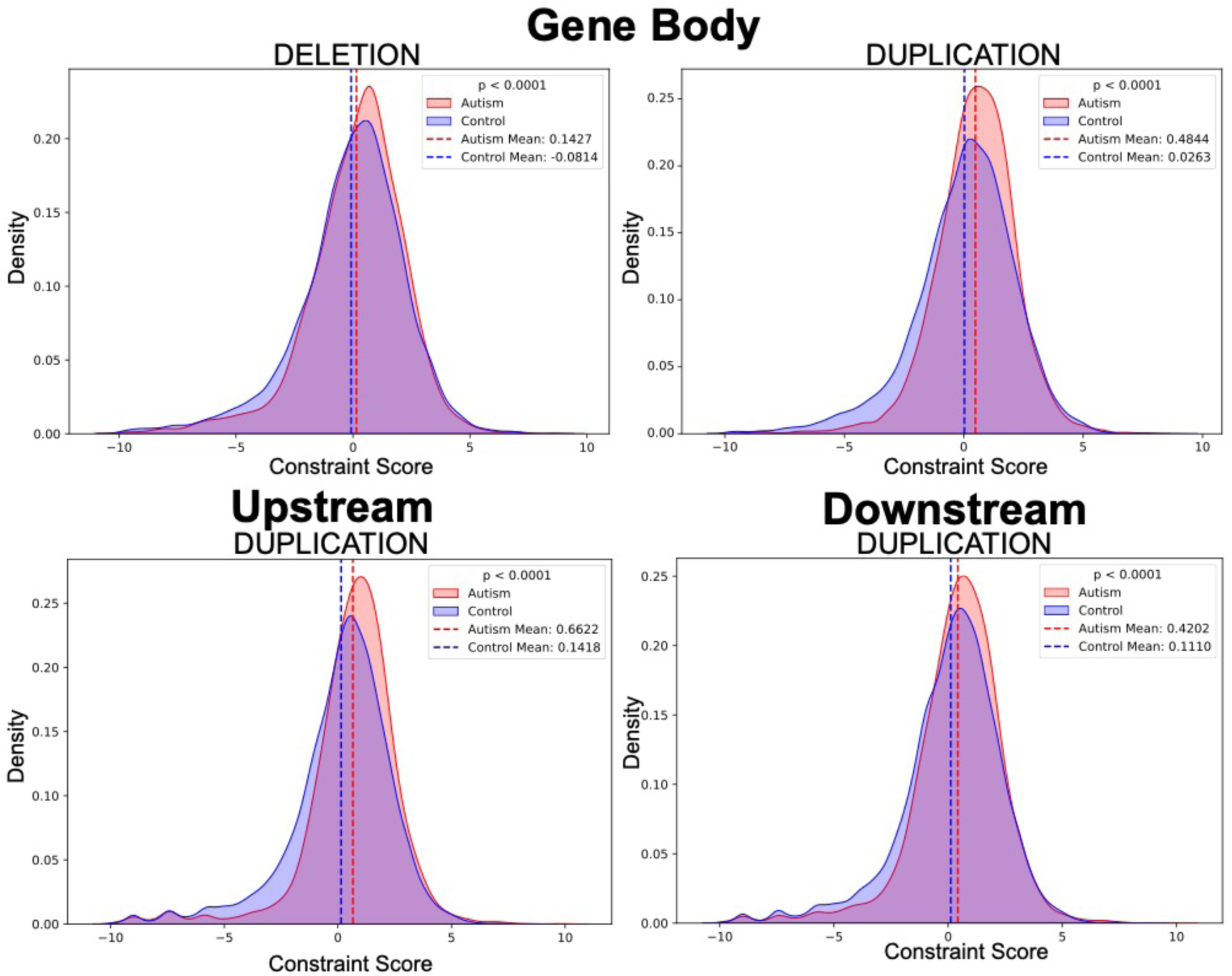
Duplication constraints are significantly different between Autism cases and controls inside and outside of the gene body. Four density graphs representing two SV types with significantly different constraint scores across Autism-control groups and gene regions. The top two graphs show significantly different constraints across groups for the gene body. The bottom two show up-stream and down-stream differences.

We then focused on DUPs in the top quartile of PGS gene weight and observed a significant enrichment in rSVs overlapping higher constraint regions in Autism compared to controls (p = 0.001, Autism z mean = 0.40, control z mean = −0.66) (**Figure 6**). This trend was not observed for genes in the bottom quartile of PGS effect (p = 0.14, case z mean = 1.0, control z mean = 0.54) (**Figure 6**) or across other SV types. In summary, DUPs were found to overlap higher constraint windows more frequently in the Autism group compared to controls for SVs overlapping, or proximal to, large-effect autism PGS genes.

**Figure 6:**
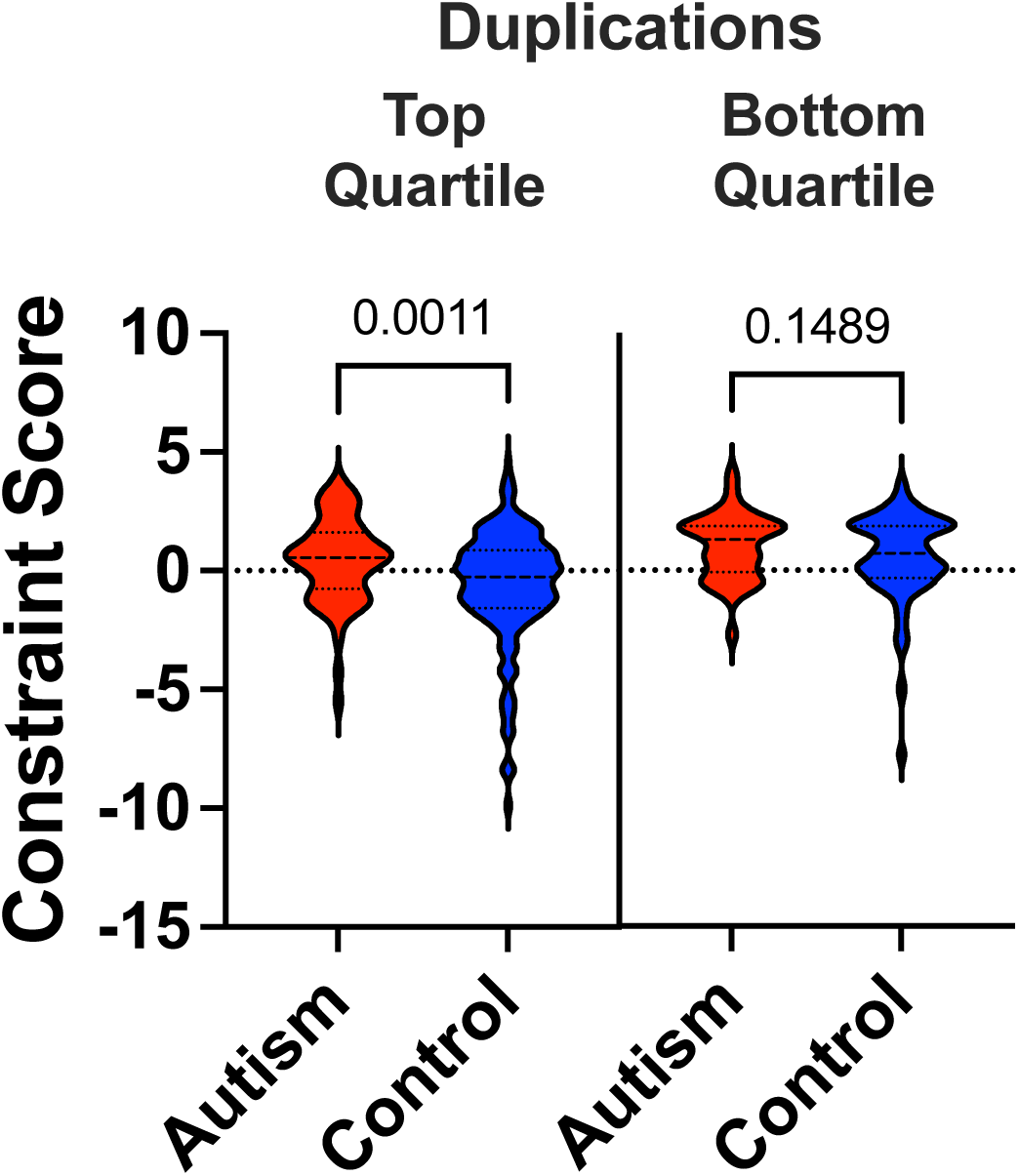
Constraint scores for duplications in top PGS quartile genes are significantly different across Autism cases and controls. Violin plot of Autism and control duplication constraint scores within top and bottom quartile genes.

## DISCUSSION

Our study highlights the significance of rare structural variants (rSVs) in complex rare diseases such as Autism. By utilizing long-read PacBio HiFi sequencing, we have enhanced the detection and characterization of rSVs in a rare disease cohort, particularly deletions (DELs), duplications (DUPs), and insertions (INSs) observed within or proximal to autism genes. Our analysis indicates a higher incidence of INSs and DELs in genes linked to Autism, with DUPs occurring frequently in large-effect PGS genes with high genomic constraint, suggesting a potential pathogenic influence. The integration of PGS has provided further insight into the genetic predisposition of various phenotypes within the cohort by using correlations between PGS and the clinical presentation of Autism.

Large-scale lrWGS assays in a rare disease cohort such as GA4K enables insights into structural variation not previously explored. This analysis identified specific genes with significant rSV intersections in the context of Autism, such as INSs in the gene body of CTNNA3, the upstream region of CDH2, and downstream of SOX5. Interestingly, all three genes were not only associated with the PGS for Autism, but they were also found in autism gene databases (Banerjee-Basu and Packer 2010; Pereanu et al. 2018). Their regional frequencies were complex, where the flanking enrichments of CDH2 and SOX5 were seen in over 63% of the Autism group, and only 42% of the group was enriched in the body of CTNNA3. This suggests that flanking enrichments of previously correlated Autism genes may provide important insights into the pathogenic influence of these genes on associated disease phenotypes. Additionally evaluating rSVs in genes with investigative association to Autism, we further resolved rSV enriched genes in Autism individuals as autistic-disorder- and cytoskeleton-specific through gene ontology enrichment. Furthermore, the online database of Autism spectrum disorder genes by the Simons Foundation Autism Research Initiative (SFARI) lists 1,176 genes related to the disorder through SNVs (Abrahams et al. 2013). In our study, we implicated over 5,000 genes using PGS associations and resolved 4,271 of those genes with rSV overlaps in our cohort. Of those 1,176 SFARI genes, 503 were in our list of intersected PGS-genes. That leaves 3,768 genes absent from SFARI but with PGS and rSV Autism overlaps identified in our cohort. Because long-read mapping-based methods for SV calling are more accurate than short-read approaches, due to their ability to provide more accurate alignments by capturing entire SV alleles in a single fragment – additionally considering that HiFi lrWGS is relatively new – our study underscores the importance in adopting lrWGS in rare disease where no single nucleotide variant is implicated.

In future studies, there should be consideration into the complexities of rare disease phenotypes and any potential comorbidities. Controlling for phenotype, age, gender, and family transmission might strengthen gene-rSV associations. Our findings underscore the complexity of the genetic factors impacting Autism risk and reinforce the importance of comprehensive analysis required in uncovering the nuances of rare disease variants. These insights lay the groundwork for more precise genetic diagnostics and the development of targeted interventions involving rSVs.

## METHODS

### Study Cohort

The study cohort described includes 497 affected probands across 419 families with 49% females and 51% males. Subjects were enrolled in the Genomics Answers for Kids (GA4K) program. Probands at age of enrollment ranged from 0 to 57 years (median = 8) (older individuals were typically ascertained as follow-up from an affected family member). Subjects were eligible for the study if they had a suspected genetic diagnosis based on clinical presentation and/or an existing molecular or cytogenic findings. The study complies with all relevant ethical regulations as approved by the Children’s Mercy Institutional Review Board (IRB) (Study #11120514). Informed or written consent was obtained from all participants prior to study inclusion. Participants were not compensated for participation.

### SV Calling

PacBio HiFi Sequencing results from GA4K proband subjects were processed through PacBio’s structural variant (PBSV, Version: 2.6.2) calling workflows after alignment to human reference GRCh38. Variants achieved a PASS score if they had at least 1 read per strand, were at least 1000 bp from the end of a contig, and away (< 1000 bp) from gaps (run of >= 50 Ns) in the reference assembly. Additionally, for PBSV, we used the following parameters: -ccs (Circular Consensus Sequence/HiFi), -m 20 (SVs larger than 20 bp were used), -A 3 (3 supporting reads were required to call a variant as homozygous alternate), -O 3 (3 supporting reads were required to call a variant as heterozygous), -P 20 (minimum phred) were used for structural variant calling. Due to known false-positives, CNVs were not included in variant analysis. Additionally, only SVs greater than 50 bp in length were used for each variant type. rSVs were defined as having a minor allele frequency (MAF) < 0.5% or allele count =< 4 in the study cohort. Somalier was used to predict ancestry (> 0.5 probability).

### GA4K-SV-FINDER

We created a python-based application for querying allele frequencies and genes associated with SVs in our cohort. This application can be found, along with a readme and the raw data file, in a GitHub repository available at https://github.com/smail-lab-cmh/ga4k-sv-finder.

### Polygenic Risk Scores

Polygenic risk scores were calculated for each specified polygenic score using open-source PGS available in the PGS catalog (www.pgscatalog.org). Associated PGS variant coordinates were downloaded from their respective catalog entries. PGS variants were converted to hg38 coordinates using dbSNP26 (version 155) where necessary. PGS variants were restricted to autosomes only and variants mapping to the HLA region were removed. Individual-level PGS were calculated using PLINK (version 1.9) (Purcell et al. 2007) (“sum” flag) on all variants available in the GA4K imputed genotype callset with R2>=0.8 (“exclude-if-info” flag). PGS scores were converted to Z-scores within each PGS in the full GA4K EUR ancestry cohort (proband and other available family members, N = 7,436). Individuals with an extreme outlier PGS Z-score (abs(PGS Z-score)>=10) in one or more PGS were removed from the final PGS dataset.

### Intersect

To create compatible chromosomal coordinate files for structural and PGS variants, pseudo-BED files were first created for each PGS SNP coordinate file, containing the chromosome, coordinates, base change, and associated weight within the PGS as a z-score. This file was first intersected, using the bedtools intersect command (v2.31.0), with a gene position BED file (gencode v26 hg38) to determine the gene the SNP was associated with. After determining the genes associated with the PGS, they were then intersected with pseudo-BED files generated from each pbsv VCF from the cohort. This created intersect files that provide the chromosomal coordinates for the SV and the genes, which included sv type, length, weight, and overlap length.

### Flanking Regions

Both up- and down-stream intersects were performed by first using BEDtools Slop command on gene start and end coordinates separately. For example, start coordinates are transformed into a 200kb upstream value, expanding the gene start coordinate within the boundaries of its respective chromosome. When intersecting these expanded genes with SVs, any SV found to enrich the gene body that is also matching an SV found in the new flanking region is removed from the flanking region analysis, as it was already intersecting the gene body. In doing this, we separated SVs found up- or down-stream of the gene from the SVs overlapping the gene body itself.

### Gene Weighting

Gene coordinates were retrieved from the gencode v26 hg38 basic annotation file (gencodegenes.org). Gene coordinates were intersected by weighted PGS variant coordinates. For each gene, PGS variant weights were transformed to absolute values before taking the maximum value across all intersecting variants. In turn, this provided an absolute maximum weight per gene. Following gene weighting, gene coordinates were intersected by rSVs.

### Constraint Scoring

Each rSV was intersected with the gnomAD v3 QCed genomic constraint by 1kb regions file (file name: constraint_z_genome_1kb.qc.download.txt.gz). This file scores 1kb regions throughout the genome with a constraint z-score value. Each of the proband rSVs were intersected with these 1kb constraint windows. For window density assessment of each rSV type, all intersected constraint windows are considered. For top and bottom quartile assessment, the scores are averaged across windows for each SV, excluding non-coding regions or any undefined region lacking a constraint value. This produces a single score for each SV intersecting a gene.

### Statistical and Enrichment Analysis

Graphing and statistics were generated using Graph Pad Prism 9 and Python (SciPy-Stats & Pandas). For enrichment, SV frequencies between Autism and control groups were determined and Chi-Square tests were used to determine significant differences. When SV counts were low for a gene (<5), a Fisher’s Exact test was used. These tests were performed for body, upstream, and downstream of genes separately.

### Gene Analysis

Gene analysis was conducted using the DISEASES database, AutDB, GeneCards Version 5.19, and Enrichr (Grissa et al. 2022; Stelzer et al. 2016; Chen et al. 2013; Pereanu et al. 2018).

## ADDITIONAL INFORMATION

Further information and requests for resources should be directed to and will be fulfilled by the corresponding author, Cas LeMaster.

## DATA AVAILABILITY

GA4K study data can be found at the ANVIL host at https://anvilproject.org/data/studies/phs002206/workspaces. Code used, as well as the application for querying SVs (GA4K-SV-FINDER) can be found in a git-hub repository at https://github.com/smail-lab-cmh/ga4k-sv-finder.

## ACKNOWLEDGEMENTS

C.S. is supported by NIH grant R35GM146966. We thank all the individuals that participated in making the GA4K study possible. This work was funded through internal institutional funds from Children’s Mercy Research Institute and Children’s Mercy Kansas City.

## AUTHOR CONTRIBUTIONS

Conceptualization: C.S., T.P, C.L.; data analysis: C.L., C.S., C.S-S., B.G., W.C., J.J.; writing: C.L., C.S.; R.M.; editing: all authors; funding: C.S.

