## Supplementary figures and images for "Mapping structural variants to rare disease genes using long-read whole genome sequencing and trait-relevant polygenic scores"

### Supplemental Figure 1

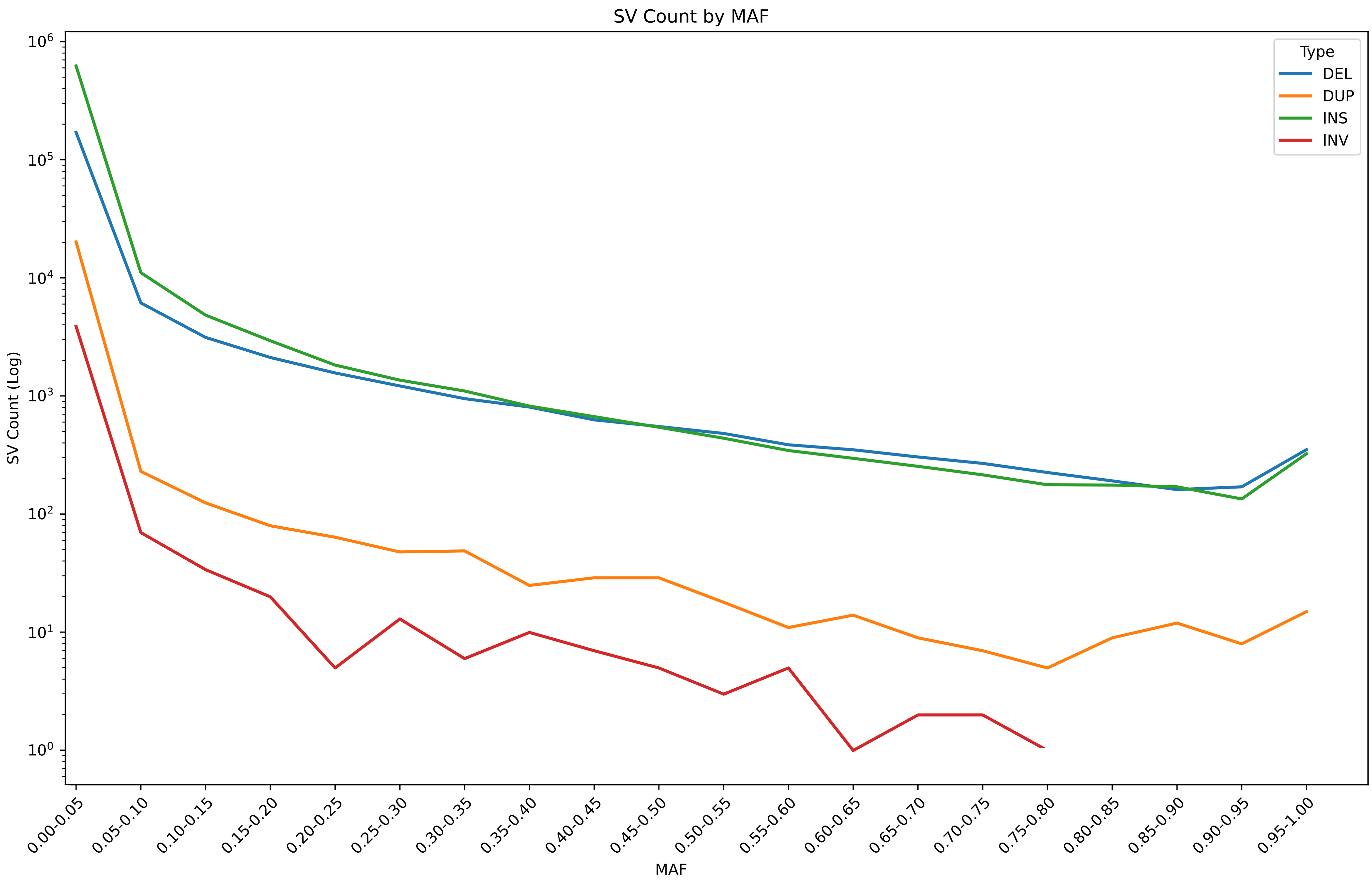
